# Performance of six SARS-CoV-2 immunoassays in comparison with microneutralisation

**DOI:** 10.1101/2020.05.18.20101618

**Authors:** AJ Jääskeläinen, S Kuivanen, E Kekäläinen, MJ Ahava, R Loginov, H Kallio-Kokko, O Vapalahti, H Jarva, S Kurkela, M Lappalainen

**Author notes:** equal contribution (S Kurkela, M Lappalainen). **Corresponding author:** Anne J. Jääskeläinen **Email:** **Address:** Helsinki University Hospital, HUSLAB, Virology and Immunology P.O.B. 720 (Topeliuksenkatu 32) FIN-00029 HUS, Finland.

## Abstract

There is an urgent need for reliable high-throughput serological assays for the management of the ongoing COVID–19 pandemic. Preferably, the performance of serological tests for a novel virus should be determined with clinical specimens against a gold standard, i.e. virus neutralisation.

We evaluated specificity and sensitivity of six commercial immunoassays for the detection of SARS-CoV-2 IgG, IgA and IgM antibodies, including four automated assays [Abbott SARS-COV-2 IgG (CE marked), Diasorin Liaison^®^ SARS-CoV-2 S1/S2 IgG (research use only), and Euroimmun SARS-CoV-2 IgG and IgA (CE marked)], and two rapid lateral flow (immunocromatographic) tests [Acro Biotech 2019-nCoV IgG/IgM (CE marked) and Xiamen Biotime Biotechnology SARS-CoV-2 IgG/IgM (CE marked)] in comparison with a microneutralisation test (MNT). Two specimen panels from serum samples sent to Helsinki University Hospital Laboratory (HUSLAB) were compiled: the patient panel included sera from PCR confirmed COVID–19 patients, and the negative panel included sera sent for screening of autoimmune diseases and respiratory virus antibodies in 2018 and 2019. The MNT was carried out for all COVID–19 samples (70 serum samples, 62 individuals) and for 53 samples from the negative panel. Forty-one out of 62 COVID–19 patients showed neutralising antibodies with median of 11 days (range 3–51) after onset of symptoms.

The specificity and sensitivity values of the commercial tests against MNT, respectively, were as follows: 95.1%/80.5% (Abbott Architect SARS-CoV-2 IgG), 94.9%/43.8% (Diasorin Liaison SARS-CoV-2 IgG), 68.3%/87.8% (Euroimmun SARS-CoV-2 IgA), 86.6%/70.7% (Euroimmun SARS-CoV-2 IgG), 74.4%/56.1% (Acro 2019-nCoV IgG), 69.5%/46.3% (Acro 2019-nCoV IgM), 97.5%/71.9% (Xiamen Biotime SARS-CoV-2 IgG), and 88.8%/81.3% (Xiamen Biotime SARSCoV-2 IgM). This study shows variable performance values. Laboratories should carefully consider their testing process, such as a two-tier approach, in order to optimize the overall performance of SARS-CoV-2 serodiagnostics.

## Introduction

Serosurveys are considered essential for creating timely snapshots for global and regional public health management of the ongoing COVID–19 pandemic (Winter and Hegde, 2020). Thus, there is an urgent need for the development of high-throughput serological assays, which enable population screening, as well as other epidemiological investigations.

Setting up a serological assay for a completely novel pathogen is challenging in many respects. At present, there is inadequate knowledge as to when and what kind of immune response follows SARS-CoV-2 infection (Okba et al. 2020) We are also yet to learn about factors that may disturb reliable serology, such as potential cross reaction from seasonal coronaviruses.

The aim of this study was to evaluate the performance of four automated immunoassays for SARS-CoV-2 serology from internationally recognized providers: Abbott, Diasorin, and Euroimmun. In addition, we evaluated two rapid lateral flow assays. The evaluation of Abbott SARS-COV-2 IgG (CE marked), Diasorin Liaison^®^ SARS-CoV-2 S1/S2 IgG (research use only), and Euroimmun SARS-CoV-2 IgG and IgA (CE marked), and two rapid lateral flow (immunocromatographic) tests of Acro Biotech 2019-nCoV IgG/IgM (CE marked) and Xiamen Biotime Biotechnology SARSCoV-2 IgG/IgM (CE marked), was conducted against a SARS-CoV-2 microneutralisation test (MNT) by using clinical serum specimens.

## Materials and methods

The patient samples consisted of serum specimens sent to the Department of Virology and Immunology, Helsinki University Hospital Laboratory, Finland for diagnostic purposes. A subset of these specimens has been included in a previous publication evaluating the Euroimmun SARS-CoV-2 IgG and IgA assays, and are included here for comparison (Jääskeläinen et al. 2020).

### Serum samples comprising the negative panel

The negative panel consisted of 81 serum samples (from 81 individuals) (median age 64 years, range 2–89 years; 33 males, 31 females). All of these samples originated from 2018–2019, i.e. before the circulation of SARS-CoV-2 in Europe.

Thirty-nine out of 81 samples contained autoantibodies: 21 had anti-nuclear antibodies in Hep-2 cell IFA analysis [NOVA Lite® DAPI ANA Kit, Inova Diagnostics, California, USA; staining patterns: homogenous, 7/21; speckled 7/21; centromere 4/21; centromere+anti-mitochondrial antibodies 1/21; nucleolar 1/21; speckled and nuclear dots 1/21], 10 were positive for phospholipase receptor A2 (PLA2R) antibodies (Anti-Phospholipase A2 Receptor IIFT (IgG) (Euroimmun, Lübeck, Germany), three for glomerular basement membrane (GBM) antibodies (EUROPLUS kidney (monkey) and GBM antigen IIFT, Euroimmun, Lübeck, Germany), and five for antineutrophil cytoplasmic (ANCA) antibodies (NOVA Lite® ANCA IFA Kit (Inova Diagnostics, California, USA) (Table 1). Presence of rheumatoid factor was tested for these 39 samples using RapiTex® RF (Siemens Healthcare Diagnostics, Erlangen, Germany) agglutination assay.

**Table 1.**
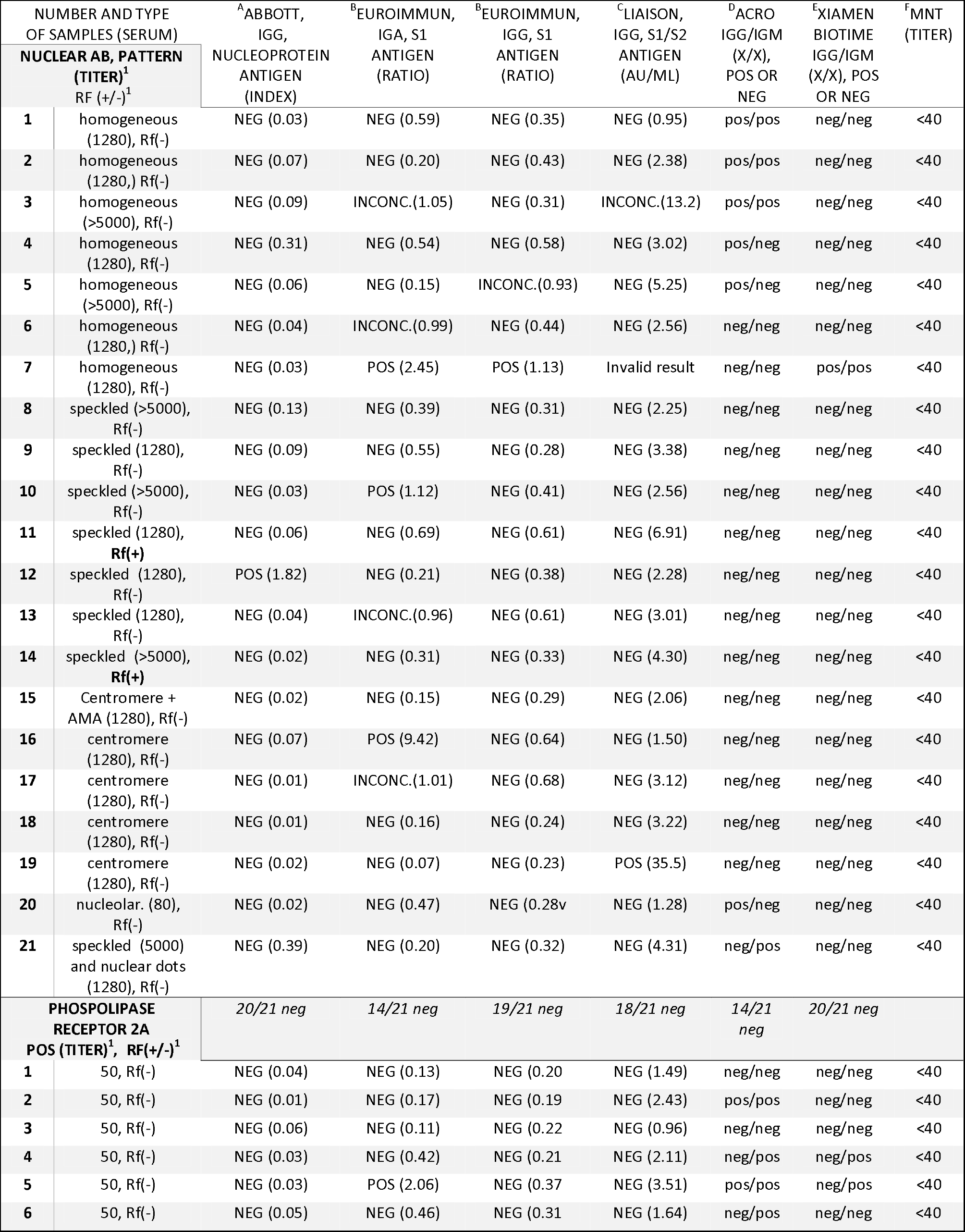
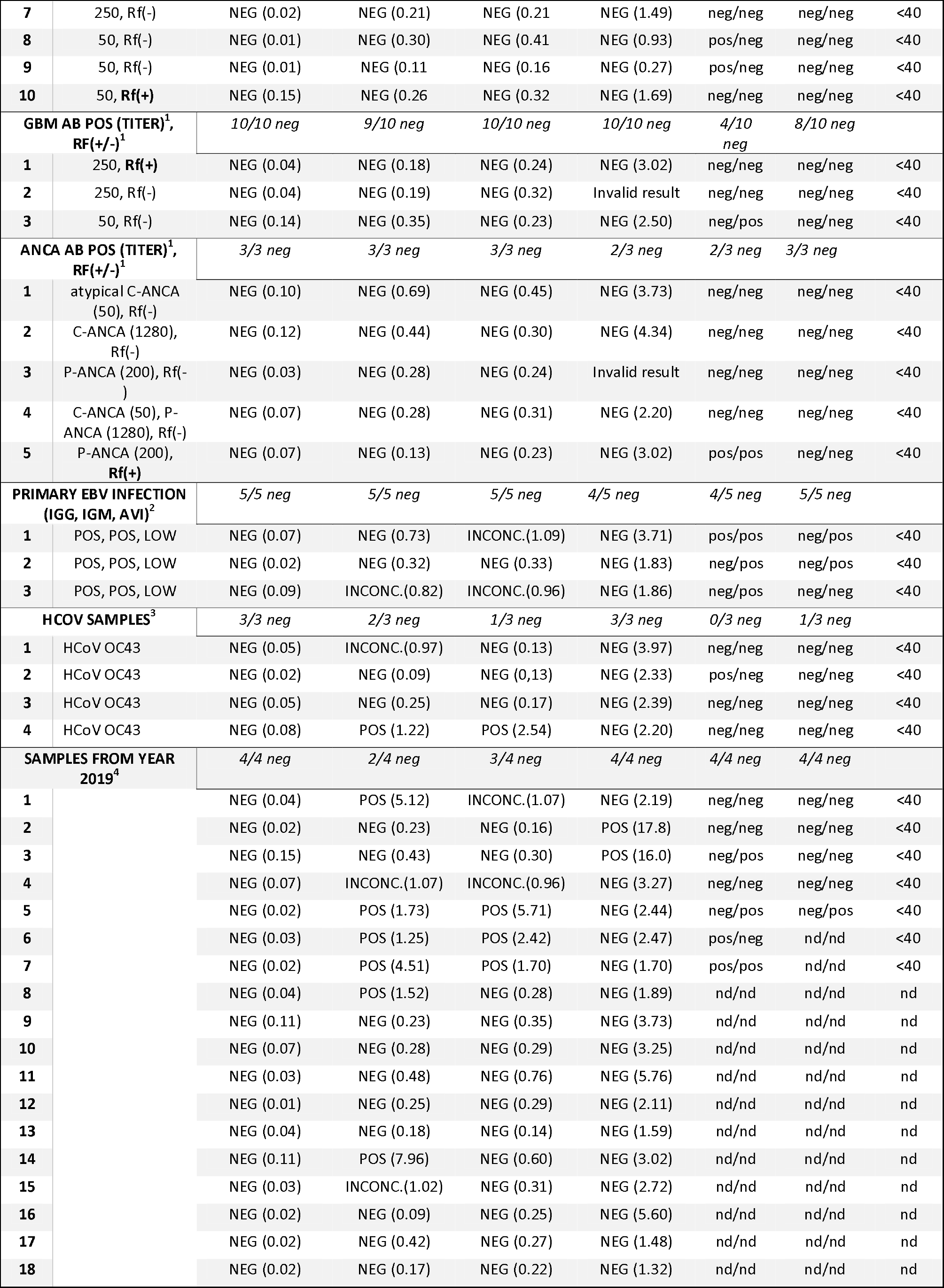
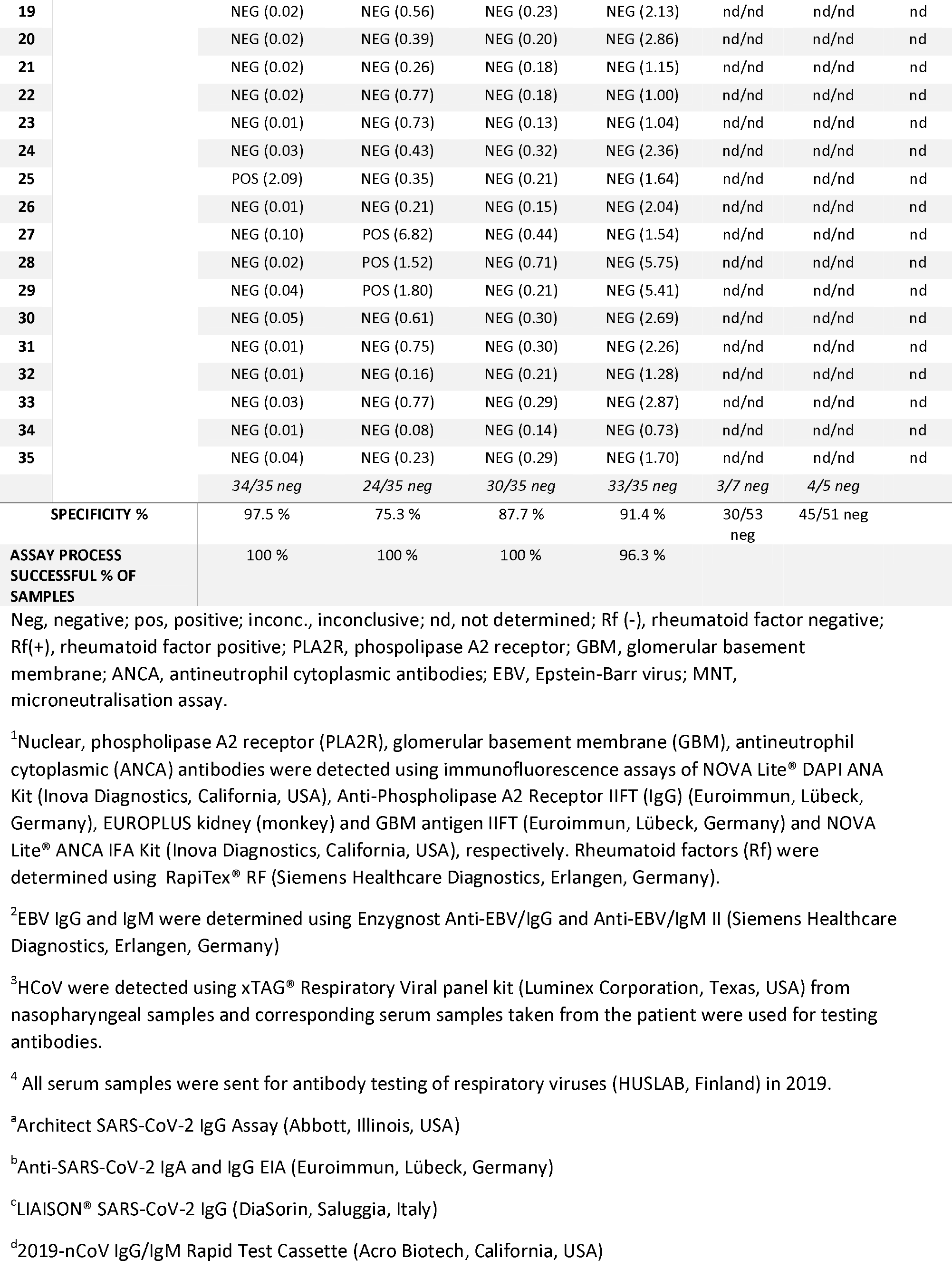
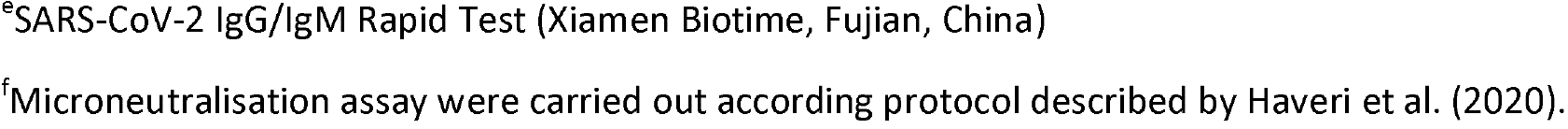
Negative serum sample panel consisting of samples collected retrospectively during years 2018–2019, prior the SARS-CoV-2 epidemic.

Three serum samples were from patients with primary Epstein-Barr virus infection (mononucleosis; EBV IgG and IgM were determined using Enzygnost Anti-EBV/IgG and Anti-EBV/IgM II (Siemens Healthcare Diagnostics, Erlangen, Germany)), four were from patients who had an ongoing Human coronavirus (HCoV) OC43 infection. HCoVs were detected using xTAG® Respiratory Viral panel kit (Luminex Corporation, Texas, USA) from nasopharyngeal samples and corresponding serum samples were collected and used for testing antibodies. In addition, 35 were serum samples originally sent for testing of respiratory virus antibodies (Helsinki University Hospital Laboratory, HUSLAB, Helsinki, Finland; Table 1).

### Serum samples comprising the COVID–19 patient panel

The patient panel consisted of serum samples from coronavirus 19 disease (COVID–19) patients, who had been diagnosed by PCR-based methods from nasopharyngeal samples in our laboratory. For molecular testing, three different methods were used: cobas® SARS-CoV-2 test on the Cobas® 6800 system (Roche Diagnostics, Basel, Switzerland), Amplidiag® COVID–19 test (Mobidiag, Espoo, Finland) and a protocol based on Corman et al (2020).

In total, 70 serum samples from 62 individuals (median age 54 years, range 24–86 years; 28 males, 34 females; Table 2) were available for this study. Data were collected and samples treated according to permit HUS/32/2018 (Helsinki University Hospital, Finland).

**Table 2.**
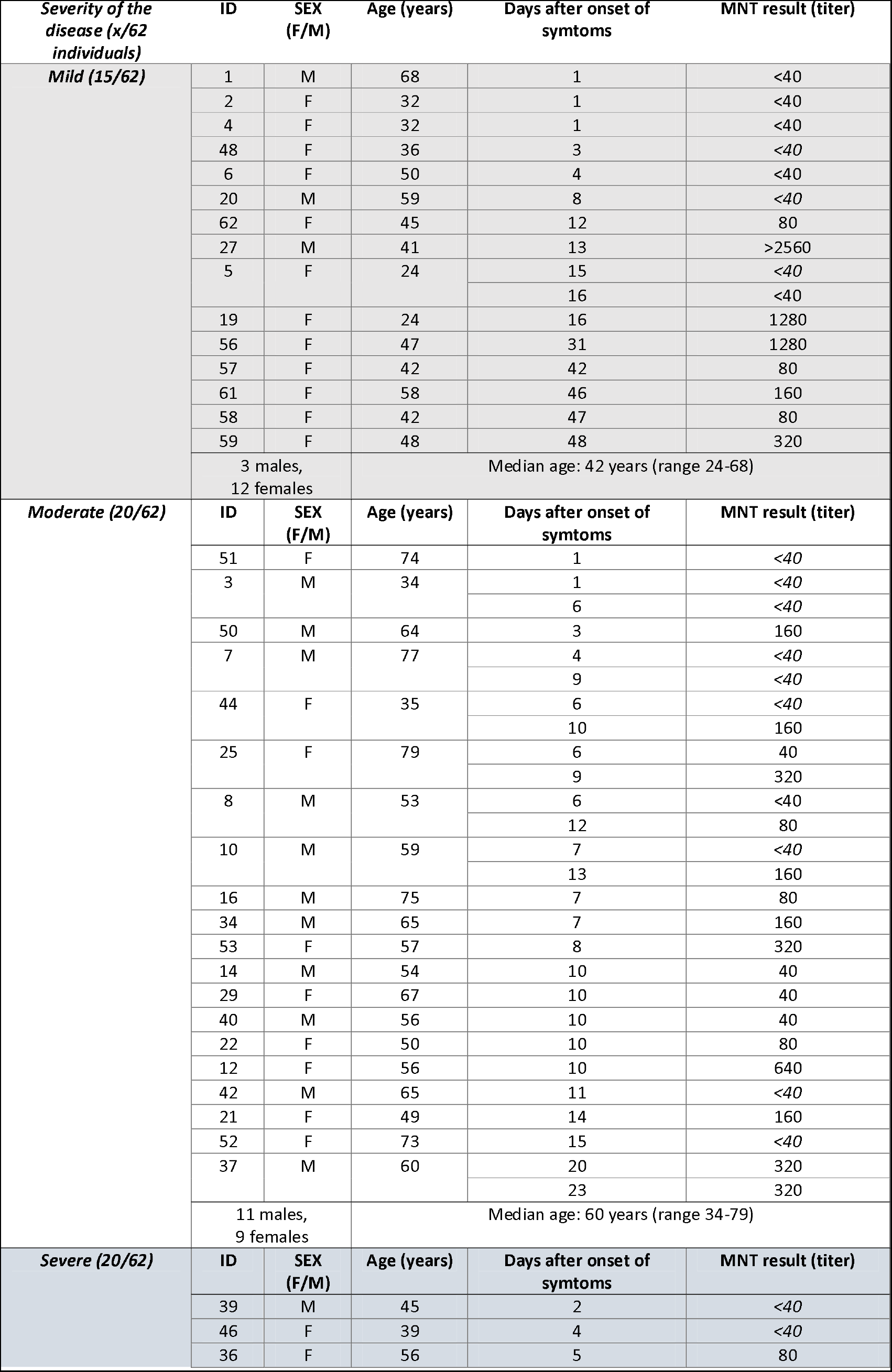
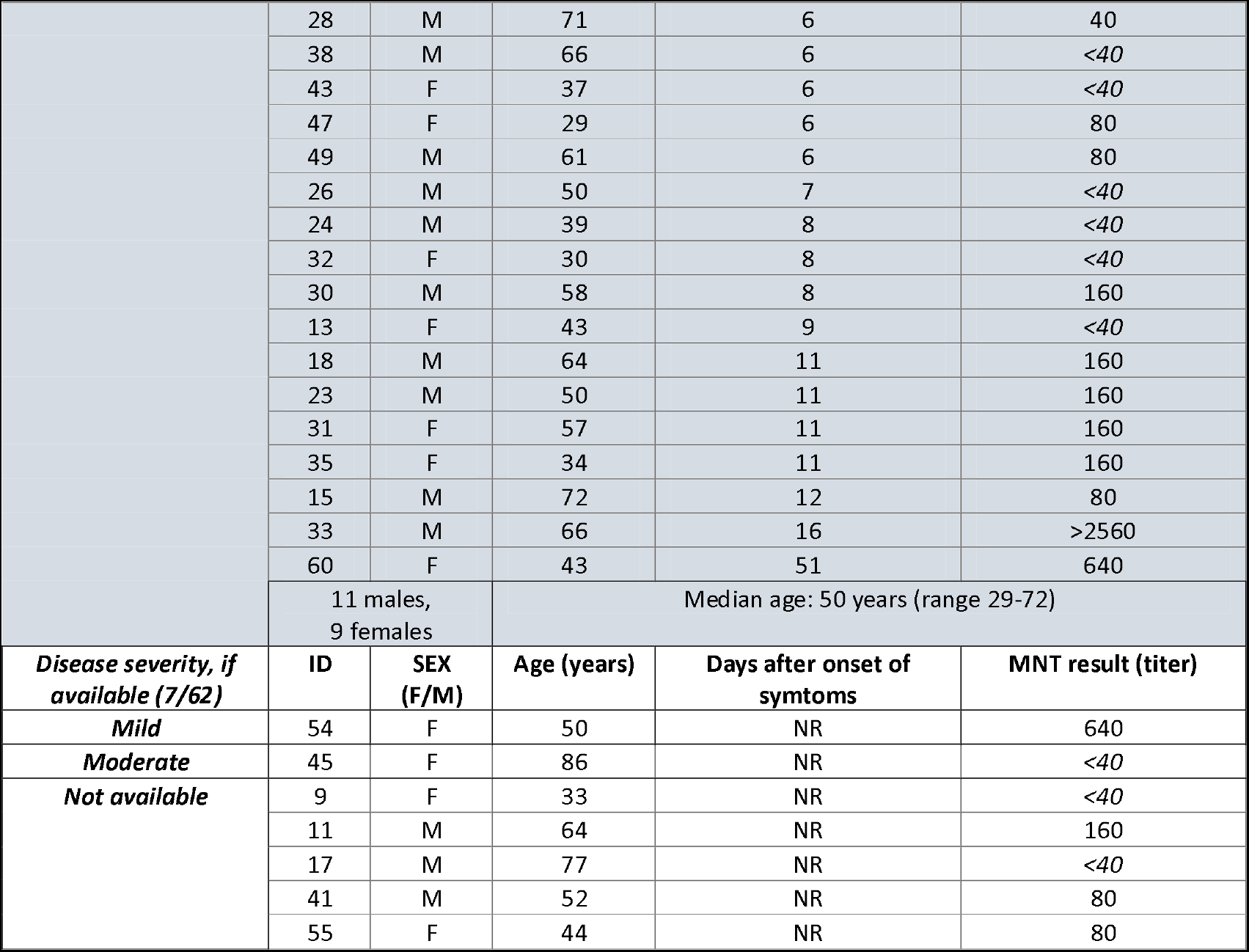
Demographic data and MNT results of all 62 COVID–19 patients.

| *Severity of the disease (x/62 individuals)* | ID | SEX (F/M) | Age (years) | Days after onset of symptoms | MNT result (titer) |
| --- | --- | --- | --- | --- | --- |
| **Mild (15/62)** | 1 | M | 68 | 1 | <40 |
|  | 2 | F | 32 | 1 | <40 |
|  | 4 | F | 32 | 1 | <40 |
|  | 48 | F | 36 | 3 | <40 |
|  | 6 | F | 50 | 4 | <40 |
|  | 20 | M | 59 | 8 | <40 |
|  | 62 | F | 45 | 12 | 80 |
|  | 27 | M | 41 | 13 | >2560 |
|  | 5 | F | 24 | 15 | <40 |
|  |  |  |  | 16 | <40 |
|  | 19 | F | 24 | 16 | 1280 |
|  | 56 | F | 47 | 31 | 1280 |
|  | 57 | F | 42 | 42 | 80 |
|  | 61 | F | 58 | 46 | 160 |
|  | 58 | F | 42 | 47 | 80 |
|  | 59 | F | 48 | 48 | 320 |
|  | 3 males, 12 females |  | Median age: 42 years (range 24-68) |  |  |
| **Moderate (20/62)** | ID | SEX (F/M) | Age (years) | Days after onset of symptoms | MNT result (titer) |
|  | 51 | F | 74 | 1 | <40 |
|  | 3 | M | 34 | 1 | <40 |
|  |  |  |  | 6 | <40 |
|  | 50 | M | 64 | 3 | 160 |
|  | 7 | M | 77 | 4 | <40 |
|  |  |  |  | 9 | <40 |
|  | 44 | F | 35 | 6 | <40 |
|  |  |  |  | 10 | 160 |
|  | 25 | F | 79 | 6 | 40 |
|  |  |  |  | 9 | 320 |
|  | 8 | M | 53 | 6 | <40 |
|  |  |  |  | 12 | 80 |
|  | 10 | M | 59 | 7 | <40 |
|  |  |  |  | 13 | 160 |
|  | 16 | M | 75 | 7 | 80 |
|  | 34 | M | 65 | 7 | 160 |
|  | 53 | F | 57 | 8 | 320 |
|  | 14 | M | 54 | 10 | 40 |
|  | 29 | F | 67 | 10 | 40 |
|  | 40 | M | 56 | 10 | 40 |
|  | 22 | F | 50 | 10 | 80 |
|  | 12 | F | 56 | 10 | 640 |
|  | 42 | M | 65 | 11 | <40 |
|  | 21 | F | 49 | 14 | 160 |
|  | 52 | F | 73 | 15 | <40 |
|  | 37 | M | 60 | 20 | 320 |
|  |  |  |  | 23 | 320 |
|  | 11 males, 9 females |  | Median age: 60 years (range 34-79) |  |  |
| **Severe (20/62)** | ID | SEX (F/M) | Age (years) | Days after onset of symptoms | MNT result (titer) |
|  | 39 | M | 45 | 2 | <40 |
|  | 46 | F | 39 | 4 | <40 |
|  | 36 | F | 56 | 5 | 80 |

|  | 28 | M | 71 | 6 | 40 |
| --- | --- | --- | --- | --- | --- |
|  | 38 | M | 66 | 6 | <40 |
|  | 43 | F | 37 | 6 | <40 |
|  | 47 | F | 29 | 6 | 80 |
|  | 49 | M | 61 | 6 | 80 |
|  | 26 | M | 50 | 7 | <40 |
|  | 24 | M | 39 | 8 | <40 |
|  | 32 | F | 30 | 8 | <40 |
|  | 30 | M | 58 | 8 | 160 |
|  | 13 | F | 43 | 9 | <40 |
|  | 18 | M | 64 | 11 | 160 |
|  | 23 | M | 50 | 11 | 160 |
|  | 31 | F | 57 | 11 | 160 |
|  | 35 | F | 34 | 11 | 160 |
|  | 15 | M | 72 | 12 | 80 |
|  | 33 | M | 66 | 16 | >2560 |
|  | 60 | F | 43 | 51 | 640 |
|  | 11 males,<br>9 females |  | Median age: 50 years (range 29-72) |  |  |
| *Disease severity, if available (7/62)* | ID | SEX (F/M) | Age (years) | Days after onset of symptoms | MNT result (titer) |
| **Mild** | 54 | F | 50 | NR | 640 |
| **Moderate** | 45 | F | 86 | NR | <40 |
| **Not available** | 9 | F | 33 | NR | <40 |
|  | 11 | M | 64 | NR | 160 |
|  | 17 | M | 77 | NR | <40 |
|  | 41 | M | 52 | NR | 80 |
|  | 55 | F | 44 | NR | 80 |

MNT, microneutralisation assay; NR, not recorded; F, female; M, male. Disease severity based on Siddiqi et al. (2020; 6). Demographic data of 39 (IDs 1-39) out of here-presented 62 COVID-19 patients were reported in Jääskeläinen et al. (2020; 3); MNT results of all these COVID-19 patients are reported first time in this table.

### Automated immunoassays for anti-SARS-CoV-2 IgG or IgA detection

The analysis of SARS-CoV-2 IgG or IgA antibodies were carried out using the Architect Plus i2000sr Analyzer (Abbott, Illinois, USA) and SARS-COV-2 IgG kit (nucleoprotein based antigen; Abbott; CE marked), EUROLabworkstation (Euroimmun, Lübeck, Germany) and SARS-CoV-2 IgG and IgA kits (S1-based antigen; Euroimmun) and Diasorin Liaison^®^ XL (DiaSorin, Saluggia, Italy) and SARS-CoV-2 S1/S2 IgG kit (S1/S2 based antigen; DiaSorin; research use only) according to manufacturers´ instructions. All samples from the negative panel (N=81) and the patient panel (N=70) were tested with Abbott SARS-CoV-2 IgG, and Euroimmun SARS-CoV-2 IgA and IgG. All samples from the negative panel (N=81) and (due to limited kit supply) 61/70 samples (53/62 individuals) from the patient panel were tested with DiaSorin SARS-CoV-2 S1/S2 IgG.

### Rapid lateral flow tests

2019-nCoV IgG/IgM (Acro Biotech, California, USA; CE marked)] and SARS-CoV-2 IgG/IgM (Xiamen Biotime Biotechnology, Fujian, China; CE marked) rapid lateral flow (immunocromatographic) tests were evaluated. Altogether, 53/81 samples from the negative panel and all 70 specimens from the patient panel were tested with Acro Biotech, and 51/81 samples from the negative panel and 61/70 from the patient panel were tested with Xiamen Biotime.

### Microneutralisation test

MNT was conducted for 53/81 of the negative panel and all of the 70 specimens from the patient panel (Tables 1 and 2). Microneutralisation assays were carried out for 39 (39/53) samples positive for autoantibodies, three (3/53) samples from patients with primary EBV infection, four (4/53) samples from patients with acute HCoV OC43 infection, and seven (7/53) samples which were sent for testing of respiratory virus antibodies.

MNT was performed in a BSL-3 laboratory as described previously (Haveri et al.2020) with modifications. Briefly, Vero E6 cells (5⌎×⌎10^4/well) were plated on 96 w/p MEM supplemented with 10% of heat-inactivated FBS, glutamine, penicillin and streptomycin. The serum samples were heat-inactivated at 56⌎°C for 30 min and 2-fold serially diluted in triplicates starting from 1:40 in MEM supplemented with 2% of heat-inactivated FBS, glutamine, penicillin and streptomycin. Fifty plaque-forming units (PFU) of the SARS-CoV-2/Finland/1/2020 strain, passaged five times in Vero E6 cells, were added to the serum dilutions and incubated for 1 h at 37⌎°C. The growth medium was removed and the virus–serum mixture was added to the cells and incubated for 4 days at 37⌎°C with 5% CO2. Neutralising antibody levels were assessed by cytopathic effect (CPE) stained with crystal violet. The neutralisation endpoint titer was determined as the endpoint of the serum that inhibited the SARS-CoV-2 infection in at least 2 out 3 parallel wells. The MNT titer was considered as positive.

## Results

For 55 COVID–19 patients out of 62, the date of disease onset was available, and disease severity could be rated (mild, moderate or severe; based on Siddiqi et al. (2020)) (Table 2, Figure 1). In the COVID–19 patients included in this study, the earliest time point for the MNT to become positive was 3 days from onset of illness (patient ID 50), while the furthest time point for a negative MNT was 16 days from onset (patient ID 5) (Table 2). Disease severity did not appear to be reflected in the MNT titers of the patients, however, the number of patients in each category was too low to assess significance (Table 2).

**Figure 1.**
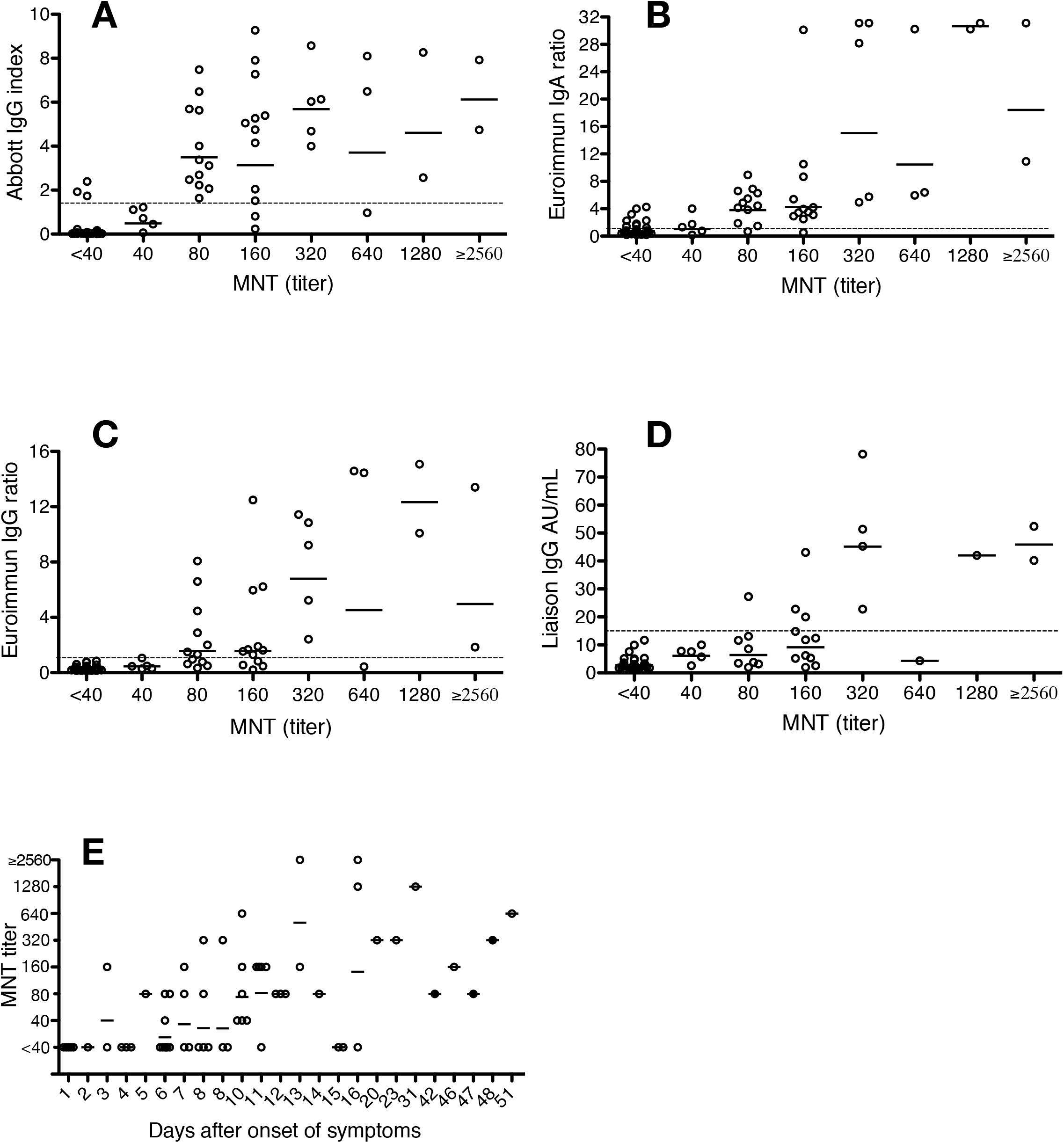
Comparison of microneutralisation test (MNT) and immunoassay results, and onset of illness. A) Abbott SARS-CoV-2 IgG assay (index); n=70. B) Euroimmun SARS-CoV-2 IgA (ratio), n=70. C) Euroimmun SARS-CoV-2 IgG (ratio), n=70, D) Liaison SARS-CoV-2 IgG (AU/mL), n=62. E) Microneutralisation titers for 63 serum samples collected from 55 COVID–19 patients, organized according to the time lapse between the onset of symptoms and the sample collection. The geometric mean is marked for each titer with a solid line and immunoassay cut-off values are indicated with a dotted line.

Numeric results of Abbott Architect SARS-CoV-2 IgG, Euroimmun SARS-CoV-2 IgA, Euroimmun SARS-CoV-2 IgG and Diasorin Liaison SARS-CoV-2 IgG were plotted against the MNT titer values (Figure 1). The geometric mean of the patient panel specimens exceeded the test cut-off at the following MNT titers: Abbott IgG (test cut-off 1.4 index exceeded with geometric mean 3.50 index at MNT titer 80); Euroimmun IgA (test cut-off 1.1 ratio exceeded with geometric mean 3.79 ratio at MNT titer 80), Euroimmun IgG (test cut-off 1.1 ratio exceeded with geometric mean 1.57 ratio at MNT titer 80), and Liaison IgG (test cut-off 15 AU/ml exceeded with geometric mean 45.1 AU/ml at MNT titer 320) (Figure 1). However, the geometric mean in the Euroimmun IgG assay lingered in close proximity (geometric mean 1.57 ratio) of the cut-off (1.1 ratio) still at MNT titer of 160.

Specificity and sensitivity for immunoassays were calculated in comparison with MNT, in which MNT titer ≥ 40 was considered positive (Table 3). Inconclusive results of the commercial assays were regarded as reactive in the performance calculations. Altogether, 53 samples from the negative panel and all of the 70 samples from the COVID–19 patient panel were tested with MNT (Tables 1–3). The specificity and sensitivity values, respectively, were as follows: 95.1%/80.5% (Abbott Architect SARS-CoV-2 IgG), 94.9%/43.8% (Diasorin Liaison SARS-CoV-2 IgG), 68.3%/87.8% (Euroimmun SARS-CoV-2 IgA), 86.6%/70.7% (Euroimmun SARS-CoV-2 IgG), 74.4%/56.1% (Acro Biotech 2019-nCoV IgG), 69.5%/46.3% (Acro Biotech 2019-nCoV IgM), 97.5%/71.9% (Xiamen Biotime SARS-CoV-2 IgG), and 88.8%/81.3% (Xiamen Biotime SARS-CoV-2 IgM). Test results from the automated immunoassays plotted against each other are shown in
Figure 2.

**Table 3.** Specificity and sensitivity of the immunoassays tested as compared to results from microneutralisation assay.

| Assay | MNT (titer) |  | Specificity<br>% (x/x) | Sensitivity<br>% (x/x) |
| --- | --- | --- | --- | --- |
|  | <40<br>(No) | ≥40<br>(No) |  |  |
| Abbott IgG+ | 4 | 33 | 95.1%<br>(78/82) | 80.5%<br>(33/41) |
| Abbott IgG inconc. | 0 | 0 |  |  |
| Abbott IgG- | 78 | 8 |  |  |
| **Total** | **82** | **41** |  |  |
| EIM IgA+ | 18 | 36 | 68.3%<br>(56/82) | 87.8%<br>(36/41) |
| EIM IgA inconc. | 8 | 0 |  |  |
| EIM IgA- | 56 | 5 |  |  |
| **Total** | **82** | **41** |  |  |
| EIM IgG+ | 5 | 26 | 86.6%<br>(71/82) | 70.7%<br>(29/41) |
| EIM IgG inconc. | 6 | 3 |  |  |
| EIM IgG- | 71 | 12 |  |  |
| **Total** | **82** | **41** |  |  |
| Liaison IgG+ | 3 | 11 | 94.9%<br>(75/79) | 43.8%<br>(14/32) |
| Liaison IgG inconc. | 1 | 3 |  |  |
| Liaison IgG- | 75 | 18 |  |  |
| **Total** | **79** | **32** |  |  |
| Acro IgG+ | 21 | 23 | 74.4%<br>(61/82) | 56.1%<br>(23/41) |
| Acro IgG- | 61 | 18 |  |  |
| **Total** | **82** | **41** |  |  |
| Acro IgM+ | 25 | 19 | 69.5%<br>(57/82) | 46.3%<br>(19/41) |
| Acro IgM- | 57 | 22 |  |  |
| **Total** | **82** | **41** |  |  |
| Xiamen Biotime IgG+ | 2 | 23 | 97.5%<br>(78/80) | 71.9%<br>(23/32) |
| Xiamen Biotime IgG- | 78 | 9 |  |  |
| **Total** | **80** | **32** |  |  |
| Xiamen Biotime IgM+ | 9 | 26 | 88.8%<br>(71/80) | 81.3%<br>(26/32) |
| Xiamen Biotime IgM- | 71 | 6 |  |  |
| **Total** | **80** | **32** |  |  |

EIM, Euroimmun; MNT, microneutralisation assay; IgG/IgA/IgM+, IgG/IgA/IgM positive; IgG/IgA/IgM-, IgG/IgA/IgM negative; inconc., inconclusive. <40 is considered negative, ≥40 is considered positive.

**Figure 2.**
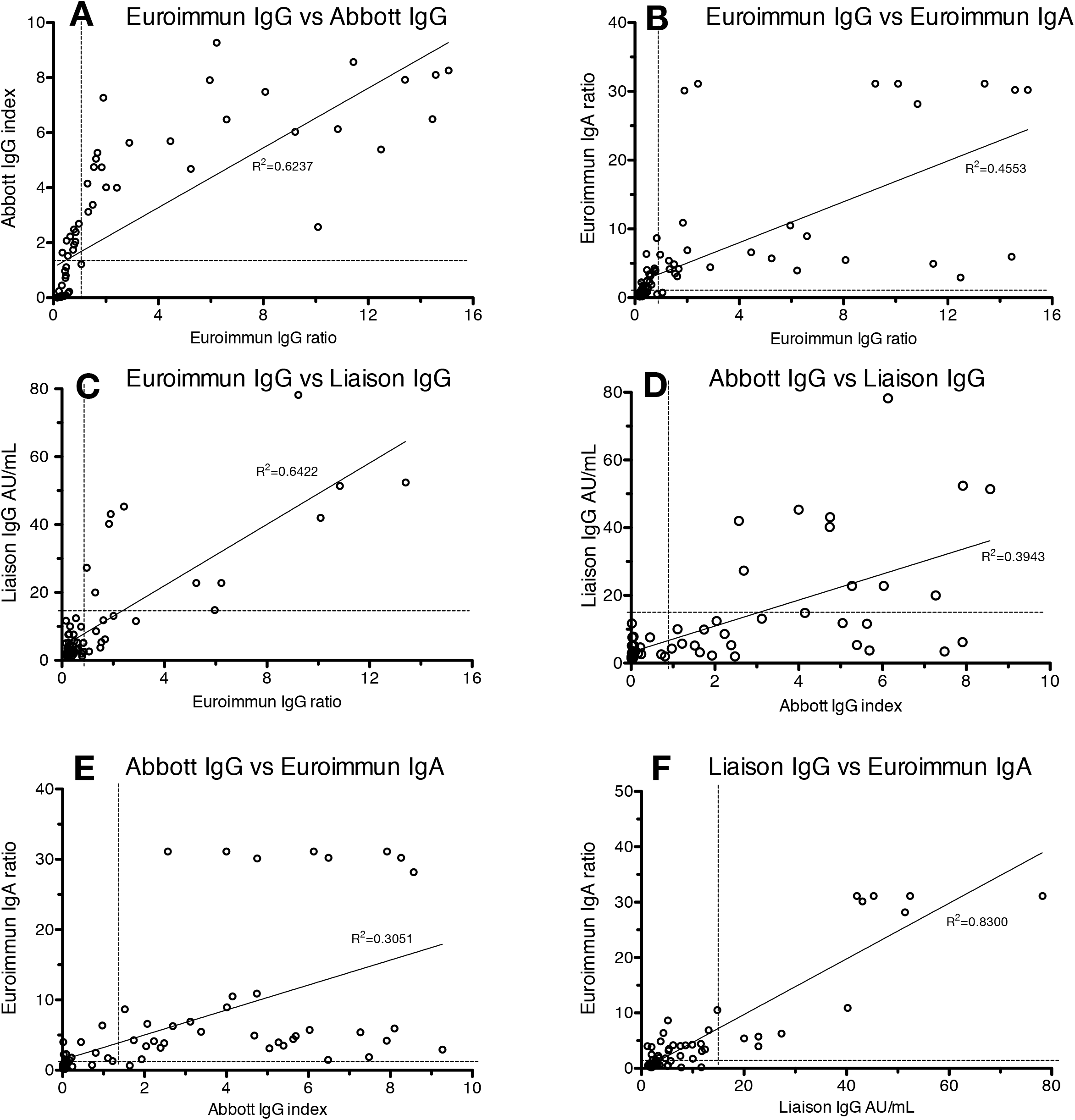
Test results from the automated immunoassays plotted against each other. A) Euroimmun IgG vs Abbott IgG, B) Euroimmun IgG vs. Euroimmun IgA, C) Euroimmun IgG vs. Liaison IgG, D) Abbott IgG vs Liaison IgG, E) Abbott IgG vs Euroimmun IgA, F) Liaison IgG vs Euroimmun IgA. The immunoassay cut-off values (dotted line) and trendlines are provided.

Rheumatoid factor was detected in five of negative panel specimens (Table 1). More detailed results are provided in Tables 1–3.

All of the six immunoassays were reactive to a varying degree of the negative panel specimens (Table 1). Particularly the Acro Biotech rapid test and Euroimmun IgA assay reacted in samples retrieved from patients with autoantibodies.

As the sensitivity of the Acro Biotech rapid test was lower than the other immunoassays tested, we randomly chose an MNT positive specimen (ID 61), conducted a dilution series of 1:2 for it, and tested the specimen again with the Acro Biotech test. An evident prozone effect was detected, and the originally negative test turned IgG positive at serum dilution 1:4 up until dilution of 1:16.

## Discussion

As serological assays for SARS-CoV-2 are now becoming available in the market in abundance (Petherick, 2020), assessment of their analytical performance by using clinical specimens is of critical importance. In this study, we assessed the specificity and sensitivity of six commercial immunoassays for the detection of SARS-CoV-2 antibodies, including two rapid lateral flow tests, in comparison with a neutralisation test. While neutralisation assays are considered to be the gold standard in terms of specificity, they also provide evidence as to development of immunity.

Eighty-one of the specimens were retrieved in 2018 and 2019 in Finland, rendering these specimens as ascertained negative for SARS-CoV-2 antibodies, and subsequently verifying the very high specificity of the neutralisation test we used (100 % were negative in MNT). We chose serum dilution 1/40 as the limit of detection for the MNT. Failure to detect very low antibody concentrations in this setup is possible. However, four of the 62 PCR-positive individuals showed neutralising antibodies without reactivity in any of the IgG tests used, suggesting a reasonable level of sensitivity in our neutralisation assay.

RF, which is an autoantibody against the Fc portion of IgG, and a common cause of cross-reactivity in immunoassays (Salonen et al. 1980), was analysed in the specimens collected in 2018 and 2019. Five out of 39 of these specimens were positive for RF; 4/5 were negative in all SARS-CoV-2 immunoassays, and 1/5 gave a positive reaction in the Acro IgG and IgM test. We conclude that the majority of positive test reactions in the six different immunoassays by using the negative serum panel from 2018–2019 were not due to RF. Of note, we observed a prozone phenomenon (Jacobs et al. 2015) by diluting specimen ID 61 for the Acro lateral flow assay. While we did not investigate prozone phenomenon extensively in this study, we do consider it may be an important cause for false negative test results. The prozone phenomenon has been reported for other lateral flow assays previously (Lee et al. 2018).

Of the automated assays included in this study, and by using the cut-off values set by the manufacturers, the best specificity values were observed with Abbott IgG (95.1%), which is in line with a previous report from the United States reporting a 99.9% specificity (Bryan et al. 2020). In our study, Liaison IgG assay (94.9%) also showed a good specificity. Euroimmun SARS-CoV-2 IgA assay had the best sensitivity (87.8%), while the sensitivity of the Liaison IgG assay was the lowest (43.8%).

When interpreting sensitivity values, the time from onset of illness in COVID–19 patients needs to be accounted for. By using the Abbott IgG assay, SARS-CoV-2 IgG seroconversion was previously reported in all patients by the day 17 post onset of illness (Bryan et al. 2020). Previous reports suggest a median seroconversion time for SARS-CoV-2 from 11 days (Zhao et al. 2020) to 13 days (Long et al. 2020). The present study also suggests a relatively long period required for serological response to take place. Even though extensive conclusions cannot be made from our data, Liaison IgG appears to turn positive at a later point in time from onset of illness in comparison with the other immunoassays evaluated in our study.

Of the two rapid lateral flow assays, the Xiamen IgG/IgM showed a good specificity (97.5% / 88.8%) with a modest sensitivity (71.9% / 81.3%). In line with a previous report (Lassaunière et al. 2020), the performance of the Acro Biotech IgG/IgM rapid test appears not to be adequate for clinical use, with specificity of 74.4% / 69.5% and sensitivity of 56.1% / 46.3%.

The currently very low seroprevalence of SARS-CoV-2 in most regions globally render low positive predictive values in the serological testing of individual patients. This can be somewhat improved by good targeting of groups tested. The analytical test performance can be optimised by placing several consecutive assays, with varying antigenic features, in the test workflow, ideally emphasizing sensitivity in the screening and specificity in the second-line testing. The very variable performance values observed in this study highlights the need for laboratories to carefully consider their testing process in order to optimize the overall performance of SARS-CoV-2 serodiagnostics.

## Data Availability

All data is available

## Acknowledgements

We would like to thank Pamela Österlund (Finnish Institute for Health and Welfare, Helsinki, Finland) for providing the virus strain. We would also like to thank (in alphabetical order) Anu Jääskeläinen (Helsinki University Hospital), Pia Jokela (Helsinki University Hospital), Laura Mannonen (Helsinki University Hospital), Tarja Sironen (University of Helsinki), Satu Suuronen (Helsinki University Hospital), Anne Toivonen (Helsinki University Hospital), and Mira Utriainen (University of Helsinki).

## References

Winter AK, Hegde ST. The important role of serology for COVID-19 control. Lancet Infect Dis 2020; pii: S1473–3099(20)30322–4.

Okba NMA, Müller MA, Li W, Wang C, GeurtsvanKessel CH, Corman VM, Lamers MM, Sikkema RS, de Bruin E, Chandler FD, Yazdanpanah Y, Le Hingrat Q, Descamps D, Houhou-Fidouh N, Reusken CBEM, Bosch BJ, Drosten C, Koopmans MPG, Haagmans BL. Severe Acute Respiratory Syndrome Coronavirus 2-Specific Antibody Responses in Coronavirus Disease 2019 Patients. Emerg Infect Dis 2020;26(7).

Jääskeläinen AJ, Kekäläinen E, Kallio-Kokko H, Mannonen L, Kortela E, Vapalahti O, Kurkela S, Lappalainen M. Evaluation of commercial and automated SARS-CoV-2 IgG and IgA ELISAs using coronavirus disease (COVID-19) patient samples. Euro Surveill. 2020;25(18):pii = 2000603.

Corman VM, Landt O, Kaiser M, Molenkamp R, Meijer A, Chu DKW, Bleicker T, Brünink S, Schneider J, Schmidt ML, Mulders DG, Haagmans BL, van der Veer B, van den Brink S, Wijsman L, Goderski G, Romette JL, Ellis J, Zambon M, Peiris M, Goossens H, Reusken C, Koopmans MP, Drosten C. Detection of 2019 novel coronavirus (2019-nCoV) by real-time RT-PCR. Euro Surveill 2020; 25(3).

Haveri A, Smura T, Kuivanen S, Österlund P, Hepojoki J, Ikonen N, Pitkäpaasi M, Blomqvist S, Rönkkö E, Kantele A, Strandin T, Kallio-Kokko H, Mannonen L, Lappalainen M, Broas M, Jiang M, Siira L, Salminen M, Puumalainen T, Sane J, Melin M, Vapalahti O, Savolainen-Copra C. Serological and molecular findings during SARS-CoV-2 infection: the first case study in Finland, January to February 2020. Euro Surveill 2020;25(11).

Siddiqi HK, Mehra MR. COVID-19 Illness in Native and Immunosuppressed States: A Clinical-Therapeutic Staging Proposal. J Heart and Lung Transplantation 2020, in press.

Petherick A. Developing antibody tests for SARS-CoV-2. Lancet 2020; 4;395:1101–1102.

Salonen EM, Vaheri A, Suni J, Wager O. Rheumatoid factor in acute viral infections: interference with determination of IgM, IgG, and IgA antibodies in an enzyme immunoassay. J Infect Dis 1980;142:250–5.

Jacobs JF, van der Molen RG, Bossuyt X, Damoiseaux J. Antigen excess in modern immunoassays: to anticipate on the unexpected. Autoimmun Rev 2015;14:160–7.

Lee G, Arthur I, Leung M. False-Negative Serum Cryptococcal Lateral Flow Assay Result Due to the Prozone Phenomenon. J Clin Microbiol 2018;56:e01878–17.

Bryan, A., Pepper, G., Wener, M. H., Fink, S. L., Morishima, C., Chaudhary, A., Jerome, K. R., Mathias, P. C., & Greninger, A. L. (2020). Performance Characteristics of the Abbott Architect SARS-CoV-2 IgG Assay and Seroprevalence in Boise, Idaho. J Clin Microbiol 2020; pii: JCM.00941–20.

Zhao J, Yuan Q, Wang H, Liu W, Liao X, Su Y, et al. Antibody responses to SARS-CoV-2 in patients of novel coronavirus disease 2019. Clin Infect Dis 2020; ciaa344

Long QX, Liu BZ, Deng HJ, Wu GC, Deng K, Chen YK, et al. Antibody responses to SARS-CoV-2 in patients with COVID-19. Nat Med 2020

Lassaunière R, Frische A, Harboe ZB, Nielsen ACY, Fomsgaard A, Krogfelt KA, et al. medRxiv preprint doi: https://doi.org/10.1101/2020.04.09.20056325

